# Transforming spontaneous premature neonatal EEG to unpaired spontaneous fetal MEG using a CycleGan learning approach

**DOI:** 10.1101/2024.03.25.24304480

**Authors:** Alban Gallard, Benoit Brebion, Katrin Sippel, Amer Zaylaa, Hubert Preissl, Sahar Moghimi, Yael Fregier, Fabrice Wallois

## Abstract

A large body of electroencephalography (EEG) studies has characterized the spontaneous neural activity of premature neonates at different gestational ages. However, evaluation of normal and pathological fetal brain development is still a challenge due to the complexity of the extraction and analysis of fetal neural activity. Fetal magnetoencephalography (fMEG) is currently the only available technique to record fetal neural activity with a time resolution equivalent to that of EEG. However, the signatures and characteristics of fetal spontaneous neural activity are still largely unknown. Benefiting from progress in machine learning and artificial intelligence, we aimed to transfer premature EEG to fMEG, to characterize the manifestation of spontaneous activity using the knowledge obtained from premature EEG.

In this study, 30 high-resolution EEG recordings from premature newborns and 44 fMEG recordings, both from 34 to 37 weeks of gestation (wGA) were used to develop a transfer function to predict the spontaneous neural activity of the fetus. After preprocessing, bursts of spontaneous activity were detected using the non-linear energy operator over both EEG and fMEG signals. Next, we proposed a CycleGAN-based model to transform the premature EEG to fMEG and vice versa and evaluated its performance with both time and frequency measurements on both forward and inverse conversions.

In the time domain, the values were similar for the mean square error (< 5%) and correlation (0.91 ± 0.05 and 0.89 ± 0.08) for the EEG to fMEG and fMEG to EEG transformations between the original data and that generated by CycleGAN. However, considering the frequency content, the CycleGAN-based model modulated the frequency content of EEG to MEG transformed signals relative to the original signals by increasing the power, on average, in all frequency bands, except for the slow delta frequency band. Our developed model showed promising potential to generate a priori signatures of fMEG manifestations related to spontaneous neural activity. Collectively, this study represents the first steps toward identifying neurobiomarkers of fetal brain development.

## Introduction

Most of our knowledge on functional neurodevelopment in humans comes from low- or high-resolution electroencephalography (EEG) of premature neonates recorded during the last trimester of gestation [1]. Fetal magnetoencephalography (fMEG), on the other hand, is the only available tool that can depict neural activity in the natural intrauterine environment and, thus, redefine the physiological and pathological functional maturation criteria of the fetus [2, 3, 4]. A considerable challenge in neurodevelopmental studies is to better understand initial cerebral development to better prevent neurodevelopmental dysfunction. It is known from decades of electroencephalography and event-related potential studies on premature neonates during the third trimester of gestation that any disturbance in the fine-tuning of the early functional wiring can have dramatic impacts on the future cognitive and sensory-motor skills in the later development of the child [5, 6, 7]. Brain development in the last trimester of gestation is an extremely dynamic process [1, 8]. Functional maturation of neuronal circuits prior to 26 to 28 weeks gestational age (wGA) relies essentially on endogenous mechanisms linked, at least in part, to spontaneous electrical activity. Subsequently, when thalamic afferents relocate from the subplate to the cortical plate, maturation mechanisms become (at least partially) sensory driven.

Given the complex nature of brain maturation dynamics, it is likely that environmental changes between ex utero and in utero, such as exposure to sound, touch, and light, as well as various physiological/neurological factors, generate singularities in the organization and communication in neural circuits and structures between the fetus and premature newborn [9, 10]. Nevertheless, as with EEG studies in premature infants, MEG studies in the fetus have demonstrated that immature neural circuits are capable of encoding exogenous information, and notably discriminating auditory and visual stimuli [11, 12]. There is also evidence for relatively similar signatures of spontaneous activity in the EEG of premature newborn and the MEG of fetus at the same gestational age (although evidence is rare in fMEG). General knowledge, supported by animal models [13, 14], indicates that the resting state activity of the early developing neural networks is characterized by discontinuity, built around bursts of activity separated by quiescent periods, called Inter-burst intervals (IBIs), of a few seconds [1, 15, 16]. The duration of the bursts and IBIs is modulated during development, and an excessively long discontinuity at a given age is an unfavorable prognostic factor. Within these bursts, specific EEG graphoelements specific to a gestational age are recorded: theta temporal activity in coalescence with slow-waves (TTA-SW) are recorded between 24 and 32 wGA, delta brushes (DB) between 28 and 36 wGA, and frontal transients (FT) between 36 and 42 wGA [1, 17]. The absence of these EEG graphoelements, which are linked to spontaneous neural activities at both cortical and subcortical levels [18, 19, 20], and that may or may not be modulated by the environment [21, 22], is also an unfavorable prognostic factor [23]. Some of these specific features (discontinuities and DB) have also been described in the fetus by fMEG recording [24]. Previous studies have used automatic techniques to detect spontaneous neural activity [25, 26]. However, such information is scarce, as no systematic studies have been performed to determine the fetal manifestations of spontaneous activity. Therefore, the general patterns of spontaneous fetal neural activity are yet to be determined.

We hypothesized that it is possible to evaluate fMEG signals using priori information derived from EEG signals of premature neonates of similar gestational age. We evaluated this hypothesis by combining two rare datasets: 30 high-resolution EEG recordings from premature newborns between 34 and 37 wGA and 44 fMEG recordings from fetuses between 34 and 37 wGA. We used our knowledge from the EEG manifestations of the spontaneous activity in premature newborns to formulate a priori hypotheses of possible manifestations of spontaneous fMEG activity. Toward this, we developed an unpaired premature EEG-to-fMEG transfer function, which was trained on our datasets corresponding to the same period of gestational age. Transferring data from one class to another has garnered significant interest in recent years with the emergence of machine learning techniques. One early approach, called Neural Style Transfer [27, 28], utilized convolutional neural networks to extract and merge the content of one image with the style of another. This method was rapidly superseded by Generative Adversarial Networks (GAN) [29], due to their ability to generate realistic data. The latter consists of an unsupervised competitive process between an artificial neural network that creates data to mimic real data and another artificial neural network that evaluates the authenticity of the generated content. Building upon this technique, CycleGAN [30] was developed to learn the mapping between two different domains, eliminating the need for paired data. The main idea behind this architecture is that the translation of data from one domain to another and then back to the original domain should result in data close to the original data. Beyond the initial use of CycleGAN for style transfer and object transfiguration in images, this technique has also been adapted to audio [31] and time-series data [32]. There are also various applications in the medical field, like electrography denoising or transfer between magnetic resonance images and positron emission-computed tomography [33, 34, 35, 36]. We adapted this approach to develop an unpaired, unbalanced transfer function between premature EEG and fMEG, enabling the extraction of spontaneous fMEG neural activity. We hypothesized that, despite the physiological and environmental factors that can influence the dynamics of these signals, there are still shared manifestations of the spontaneous neural activity between them. Given the difficulties in searching for such spontaneous activity in fMEG, this approach allows the development of templates for further automatic search engines, to be trained on fMEG. As an initial step, we aimed to estimate how an EEG signal could be rendered in fMEG using CycleGAN. This strategy was applied to bursts of activity, in addition to two main neurobiomarkers, namely DB and FT. The transfer model was developed based on short periods of bursts, under the hypothesis that these periods capture the temporal and spectral dynamics of spontaneous neural activity. The organization of this study involves, first training and validating the CycleGAN transfer function, followed by evaluating the performance on EEG to fMEG data that were not included in the training and validation phases.

## Methods

### Population

We used high-resolution EEG recorded from 30 sleeping premature neonates between 34 and 37 wGA (mean age 35.47 ± 1.09 wGA) in the neonatal intensive care unit of Amiens University Hospital (Amiens, France, Table 1). The duration of each EEG recording varied between 6.5 and 42.9 minutes, with an average of 22.9 ± 9.4 min. All infants had appropriate birth weight, size, and head circumference for their term age. They also had an APGAR score > 5 at 5 min and a normal clinical neurological assessment. None were considered to be at risk of brain damage. In particular, the results of a neurological examination at the time of the recordings had to correspond to those of the corrected gestational age, with no history of abnormal movements. The gestational age (estimated from the date of the mother’s last menstrual cycle and ultrasound measurements during pregnancy) corresponded to the degree of brain maturation (evaluated using EEG). The EEG evaluation in the follow-up at the time they left the neonatal intensive care unit had to be normal according to the EEG monitoring guidelines of the French Society of Clinical Neuroscience. One or both parents were informed about the study and provided their written informed consent. The local ethics committee (CPP Ouest I) approved the study (ID-RCB: 2021-A02556-35).

**Table 1.** Number of participants for premature high-resolution EEG and fMEG according to their gestational age at the recording date.

| Signal | wGA |  |  |  | Total |
| --- | --- | --- | --- | --- | --- |
|  | 34 | 35 | 36 | 37 |  |
| EEG | 7 | 9 | 7 | 7 | 30 |
| MEG | 13 | 11 | 12 | 8 | 44 |

We also used fMEG recorded from 44 fetuses between 34 and 37 wGA (mean age 35.34 ± 1.10 wGA) in the Tuebingen Hospital (Tuebingen, Germany, Table 1). The duration of each fMEG varied between 6 and 27 min, with an average of 22.1 ± 8.1 min. Participants were recruited from Tübingen and its surrounding areas. All mothers had uncomplicated singleton pregnancies. The recordings were approved by the local ethics committee of the Medical Faculty of the University of Tübingen and the consent to participate was signed by the mother (511/2015BO1 and 330/2010BO1). They received 10 euros per hour for their participation.

### EEG and MEG acquisition

High-resolution EEG recordings were made in the incubator, while the premature neonates were asleep, using a 64 or 124-channel HydroCel Geodesic Sensor Net with an Electrical Geodesic NetAmps 200 amplifier passing a digitized signal to Electrical Geodesics Net Station software (version 5) and were DC–50 Hz filtered. EEG was digitized at a 1000 Hz sampling rate, with a Cz vertex electrode as the reference. The electrode impedance was maintained below 5 kΩ.

The fMEG recordings were made using a SARA (SQUID Array for Reproductive Assessment, VSM MedTech Ltd., Port Coquitlam, Canada) system installed at the fMEG Center at the University of Tübingen. The system was developed to adapt the established technique of MEG to the special requirements of fetal measurements. It includes 156 primary magnetic sensors, which are distributed over a concave array, shaped to match the form of the gravid abdomen. The system also includes 29 reference sensors. Data were recorded at a sampling rate of 610.352 Hz. To attenuate the influence of external magnetic fields, the system is installed in a magnetically shielded room (Vakuumschmelze, Hanau, Germany).

### Preprocessing

The preprocessing steps for both EEG and fMEG, presented in the following subsections, are summarized in Figure 1. All steps were conducted using MNE 1.2.1 for Python 3.10.6.

**Figure 1.**
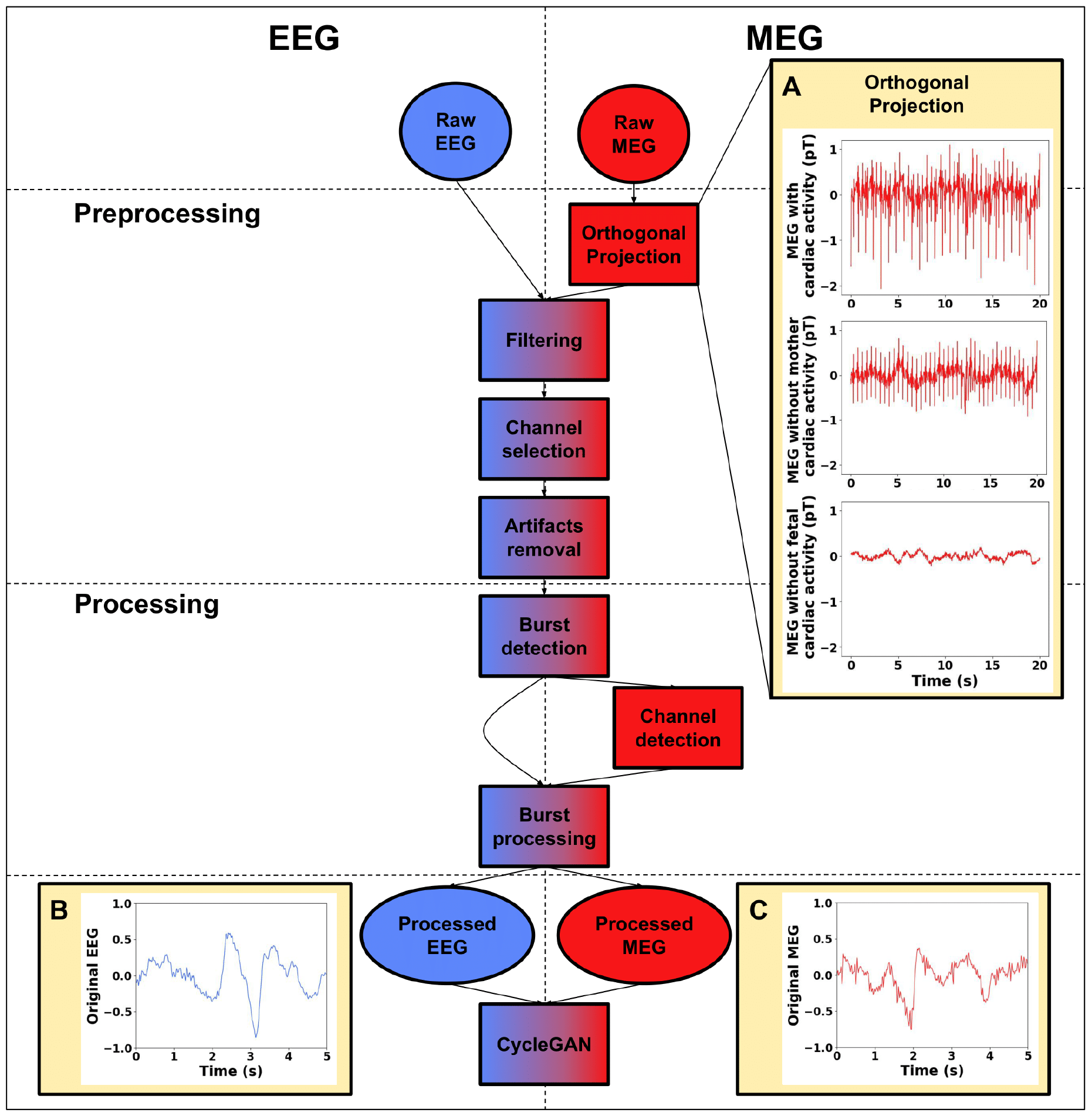
Preprocessing and processing steps for EEG (left, blue) and fMEG (right, red) signals. Circular boxes represent inputs and outputs, and rectangular boxes represent processing steps. A) Example of an orthogonal projection: top - before applying the orthogonal projection, middle - after removing the maternal cardiac artifact, and bottom - after removing the fetal cardiac artifact. Examples of normalized EEG (B) and fMEG (C) after processing.

EEG recordings were zero-phase band-pass filtered (0.5-20 Hz). Next, the spatial resolution was reduced from 64/128 channels to a low-resolution 10 bipolar-channel montage used in the clinic to fit the number of MEG channels with brain activity [37, 38, 39]. The 10 reconstructed EEG channels covered the whole head (‘Fp2-C4’, ‘Fp1-C3’, ‘C4-O2’, ‘C3-O1’, ‘Fp2-T4’, ‘Fp1-T3’, ‘T4-O2’, ‘T3-O1’, ‘Fz-Cz’, and ‘Cz-Pz’). Artifacts were removed using an amplitude threshold of 500 μV. A 0.5-s window prior to and after the detected artifacts was removed.

For MEG signals, the first step consisted of removing maternal and fetal cardiac artifacts that interfered with the measurement of brain activity (Figure 1A), using the orthogonal projection method [2, 40]. Briefly, this technique projects out physiological interference by constructing the orthogonal projection operator from signal space vectors, corresponding to the interfering components. Given the presence of two distinct interfering components (maternal and fetal cardiac activity) in the signal, the method was applied twice consecutively. The first projection minimized the cardiac artifact from the mother (Figure 1A, middle) and the second projection, that from the fetus, which was much smaller in amplitude than that of the mother (Figure 1A, bottom). The number of projectors for each step was manually selected between 1 and 3 for each phase, to achieve optimal performance (evaluated through visual inspection), while retaining what we considered as background neural activity. After cardiac artifact removal, the fMEG signals were zero-phase band-pass filtered (0.5 and 20 Hz). Finally, we conducted a visual inspection of the channels and removed any noisy ones to prevent errors in the burst-detection phase. Any remaining artifacts were removed using an amplitude threshold of 2000 fT. A 0.5-s window prior to and after the artifact was removed.

### EEG/MEG burst detection

To automatically detect bursts in premature EEG and fMEG recordings, a nonlinear energy operator (NLEO) was used [41, 42]. Briefly, to characterize the bursts, NLEO combines the amplitude and spectral content of the signal at each time point. For each sample *i* of the signal, the NLEO index was calculated as

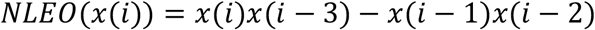

where *x* represents the signal amplitude. To calculate the NLEO index, the signal was resampled to 256 Hz. To achieve better characterization of the dynamic changes, the absolute value of the NLEO index was smoothed by calculating the average over a sliding window of 384 samples (equivalent of 1.5 s) [41]. Next, a threshold was applied to each channel, distinguishing between signals above the threshold as bursts of activity and those below as IBIs. The threshold was manually and individually optimized for each subject through visual inspection. The mean threshold values for the EEG and MEG signals were 0.81 ± 0.23 μV^2^ and 17.0 ± 6.4 fT^2^, respectively.

From previous reports [43, 44], and considering the fetal head diameter at this gestational age (∼10 cm) [45], we expected to detect the manifestations of spontaneous neural activity on only a small number of MEG channels. Therefore, a region of interest comprising 10 channels was defined based on the spatial amplitude distribution of smoothed MEG NLEO signals. The center of the region of interest was determined by the maximum amplitude of the smoothed MEG NLEO signal. Next, the nine closest channels were selected to obtain the 10 fMEG channels, likely to present signals corresponding to neural activity. The spatial distribution of the NLEO mean amplitude over the channels is illustrated in Figure 2.

**Figure 2.**
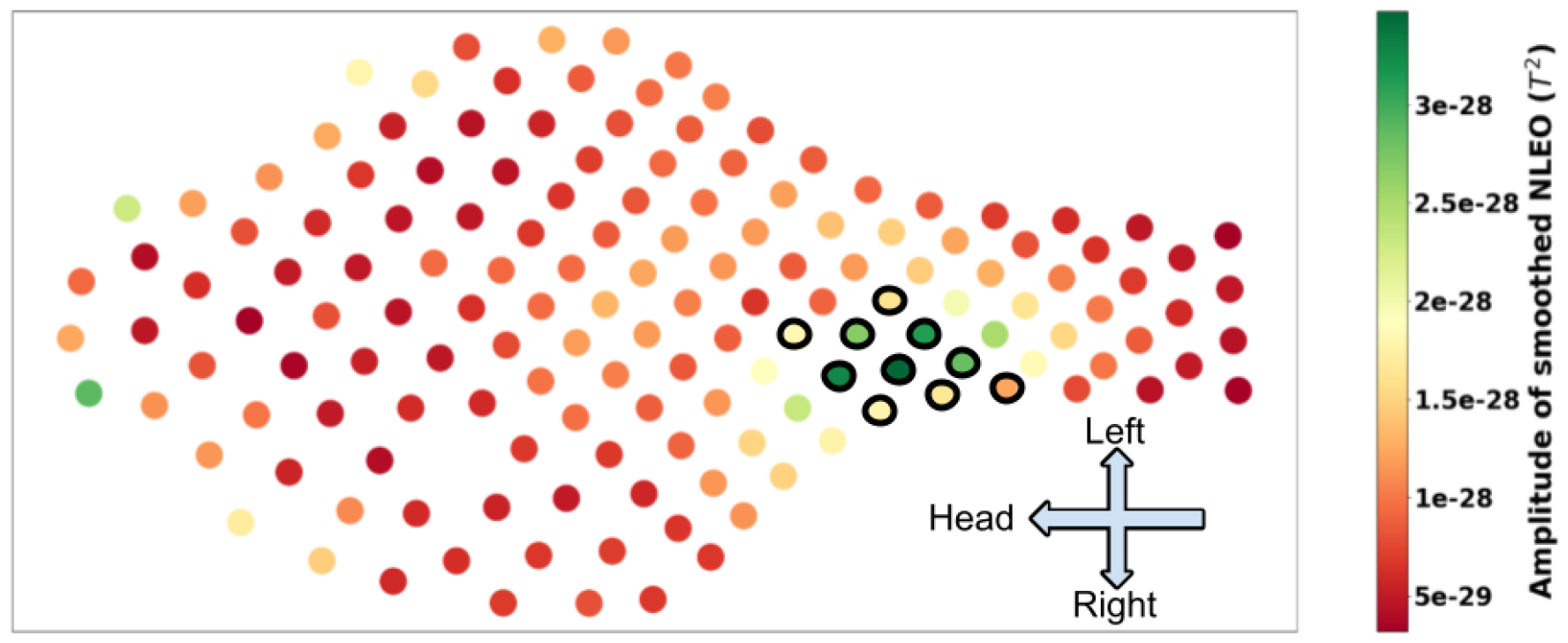
Spatial amplitude distribution of smoothed NLEO for one participant. The head of the mother is located on the left of the figure, her left on the top and her right on the bottom. The selected region of interest is shown with black circles. In this case, the centroid is localized around the center of the lower abdomen of the mother.

For both EEG and fMEG signals, a burst was finally marked if it occurred simultaneously over at least half of the electrodes/channels. Otherwise, the period was marked as an IBI. Consecutive bursts separated by an IBI shorter than 2 s were merged. Only burst periods were considered for further analyses. EEG and MEG bursts were resampled to 64 Hz to reduce the number of features provided as input to CycleGAN, while preserving data quality. The bursts of each signal were divided into 5 s segments with a 2.5-s overlap to increase the data segments for CycleGAN. Remaining bursts with a duration of less than 5 s were discarded from further analyses. The amplitude of both EEG and fMEG segments were normalized to [-1,1], using the maximum and minimum values of the whole dataset and by calculating:

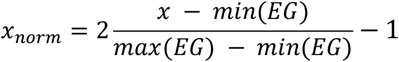

where *x*_*norm*_ is the normalized signal amplitude and *x* the signal amplitude before normalization. *min*(*EG*) and *max*(*EG*) are the minimum and maximum amplitude values of the whole 5s segment datasets (EEG or MEG). Finally, the datasets were divided into train (80%) and test (20%) subsets (Table 2).

**Table 2.** Number of EEG and MEG burst segments in the train and test subsets.

| Number of burst segments | Train | Test | Total |
| --- | --- | --- | --- |
| EEG | 8542 | 2090 | 10632 |
| MEG | 7684 | 1873 | 9557 |

### CycleGAN

CycleGAN is a machine-learning model developed for unpaired data translation, specifically image-to-image transformation from painting to real photo [30, 46]. This model is also employed in the medical field [33, 34, 35, 36]. By adapting the original model to accommodate time-series instead of images, as carried out in [32], the characteristics of this approach fit well with the case of EEG to fMEG transformation, in which the two signals do not correspond to the same participant and do not originate from the same modality, despite the fact that both provide manifestations of spontaneous neural activity at the same gestational age.

The architecture of CycleGAN used in this study is shown in Figure 3. It consists of two conditional GANs, one for data transformation from EEG to MEG and the other from MEG to EEG. Each GAN is made up of two sub-networks, the first being a generator, which takes a data item from the source domain and aims to produce a data item that is both realistic and similar to the target domain. Here, the generator consists of a convolutional neural network architecture that applies a series of convolutional and transposed convolutional layers to the input signal, transforming it into a signal from the target domain. The second sub-network is a discriminator, the aim of which is to evaluate the realism of the generated data in relation to the target domain, and thus provide feedback to the corresponding generator during the training process. This is achieved by passing a signal through a series of convolutional layers to obtain a score. The aim of the discriminator is to produce high probability scores when the input signals are the original signals and low probability scores when the input signals are produced by the generator. In addition, two main loss functions are applied. The first consists of adversarial loss (Figure 3, “Control Discriminator”), which links the generator and discriminator of the same domain. On the one hand, the generator tries to minimize loss by generating signals that are indistinguishable from the ‘real’ data, i.e., successfully ‘fooling’ the discriminator. On the other hand, the discriminator tries to maximize loss by correctly classifying the signals as ‘real’ or ‘generated’. The second loss function, specific to CycleGAN, is the cycle-consistency loss (Figure 3, “Control Cycle”). The idea is that when a signal is translated from one domain to the other, and then translated back to the initial domain, the resulting signal must be similar to the original signal. This constraint thus ensures that the content and structure of the signal are preserved during translation. During the training process, the two generators and two discriminators work in tandem. The generators aim to create convincing translations, while the discriminators strive to better distinguish between real and generated signals. Over time, through adversarial training and cycle-consistency, the generators improve their ability to generate semantically correct signals for the target domain.

**Figure 3.**
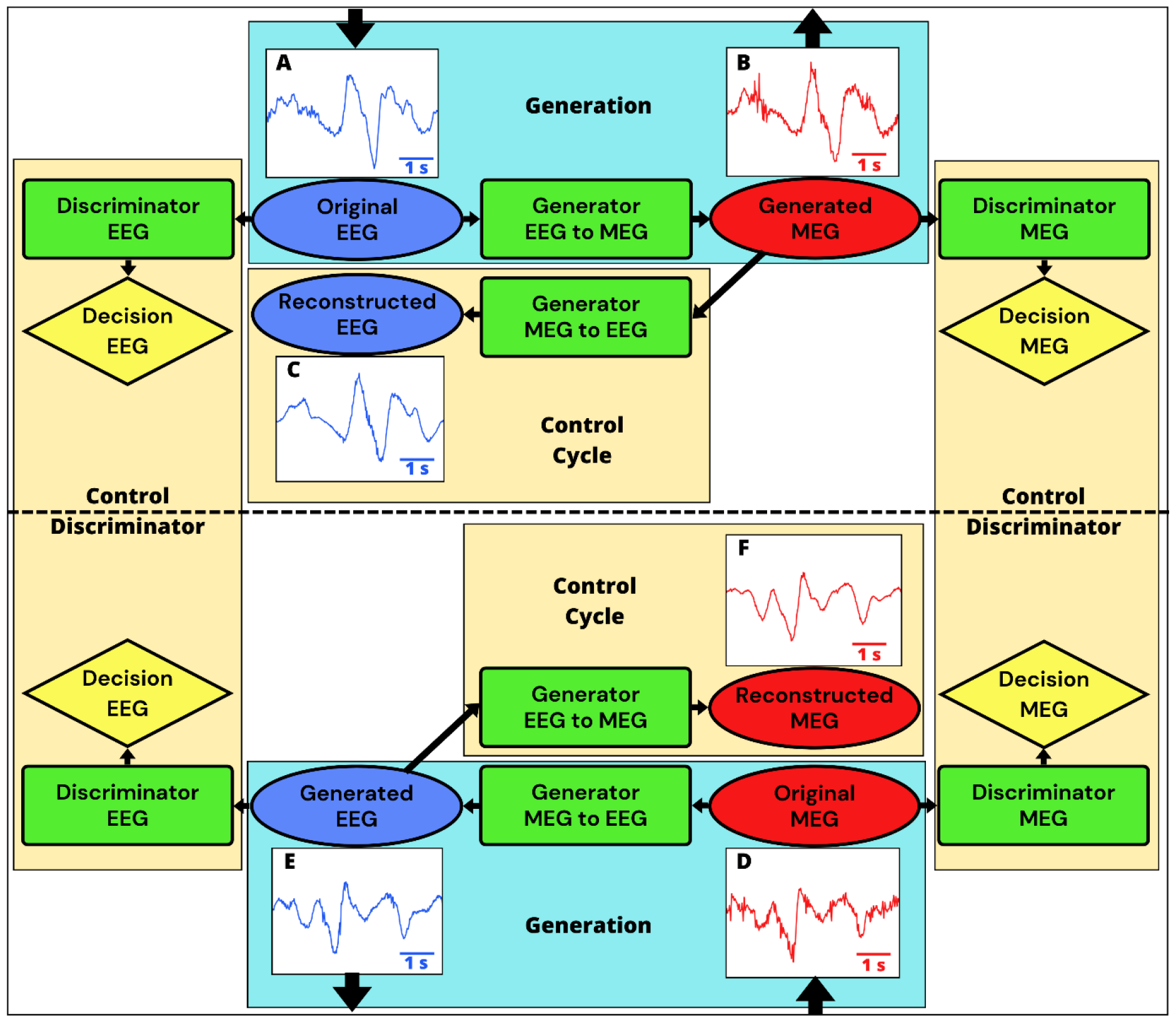
CycleGAN. The EEG signals are shown in blue, the fMEG signals in red, the neural networks in green, and the decision in yellow. The “Generation” boxes are the elements of CycleGAN that generate data from the original EEG and MEG data as input and the generated MEG and EEG data as output. The “Control Cycle” boxes correspond to the validation of the reconstruction and generation of the signal. The “Control Discriminator” boxes evaluate the ability to discriminate between the original and generated signals. A) original EEG, B) generated MEG, C) reconstructed EEG, D) original MEG, E) generated EEG, and F) reconstructed MEG.

All the calculations related to CycleGAN were conducted on the UPJV MatriCS computing platform.

### Performance evaluation

For the first step of the evaluation process, we compared the generated and reconstructed signals, for both EEG and fMEG. More precisely, we evaluated the performance of the CycleGAN-based model to generate/reconstruct data (from EEG to MEG and vice versa), using measurements from both the time and frequency domains on the test dataset to study the temporal structure and spectral content of the signals. In the time domain, we calculated the mean square error (MSE) and cross correlation between the original and reconstructed signals. In the frequency domain, we compared the frequency content of the original and reconstructed signals. Toward this, we first calculated the spectrum of each segment (reconstructed and original signals) by applying a discrete Fourier transform and producing a spectrum with a frequency resolution of 0.2 Hz. We limited the spectral analyses to the 0.5-20 Hz spectral window, as, based on the literature, the cortical manifestations of the spontaneous activity at this gestational age are mostly limited to this frequency range [1]. Next, the average spectrum was calculated for each participant. For further statistical analyses, the spectra were divided into four frequency bands, namely delta (0.5-3 Hz), theta (3-8 Hz), alpha (8-12 Hz), and lower beta (12-20 Hz). The average value of each band was calculated for each participant. A two-sample two-tailed t-test was carried out to compare the spectral content of the reconstructed and original signals at each frequency band, for both EEG and fMEG. In addition to evaluating the ability of CycleGAN to adapt to the novel data, the MSE was also calculated for the test dataset and the ratio between the mean absolute value of the original signal and the MSE was compared using a two-sample two-tailed t-test for the train and test datasets.

For the second step of the evaluation process, we studied the fMEG signals generated from the EEG signals. First, 10 clear DB and FT were selected from EEG signals for three random participants by an expert (F.W.). The EEG to fMEG CycleGAN model was then applied to the selected events. The obtained fMEG signals were visually evaluated to test the fMEG manifestations of specific spontaneous neural activity. Second, we compared the average characteristics of the EEG-based generated fMEG signals and the original fMEG signals. Toward this, we first calculated the spectral power of the original fMEG signals and the fMEG signals generated from EEG for each segment by applying a discrete Fourier transform, and then averaged over segments for each participant. The frequency resolution was 0.2 Hz and the spectral window was limited to 0.5-20 Hz, as carried out in the previous step. After division into frequency bands and averaging for each participant, a two-sample two-tailed t-test was carried out to compare the power content of each frequency band corresponding to the original fMEG and the generated fMEG signals.

## Results

### Evaluation of CycleGAN

After the training phase, EEG signals (Figure 3A) were transformed into fMEG signals (Figure 3B), using the CycleGAN model. Next, the EEG signals were reconstructed (Figure 3C) from the generated (transformed) fMEG signals. Similarly, fMEG signals (Figure 3D) were transformed into EEG signals (Figure 3E), and then reconstructed (Figure 3F). The signals reconstructed by CycleGAN were evaluated in three different ways using the test dataset: MSE, correlation, and spectral analysis. CycleGAN showed good performance for the reconstruction of both EEG and fMEG signals, with a MSE ratio of < 5% between the original and reconstructed signals in both train and test phases (Table 3). The MSE ratios (normalized to the mean absolute value of the signals) were not statistically different between the train and test datasets (t = 1.10 and p = 0.27 for EEG, t = -0.46 and p = 0.64 for fMEG). This suggests that the trained CycleGAN model was able to successfully generalize the transformation developed during the training phase to the test dataset (not seen during the training phase).

**Table 3.** The mean absolute value (calculated for each 5 s) of the original signals, the MSE values between the original and reconstructed signals, and the ratio between the mean absolute values and MSE values for both the train and test datasets for both the EEG and fMEG signals. All values are expressed as averages ± standard deviations.

|  | EEG train | EEG test | MEG train | MEG test |
| --- | --- | --- | --- | --- |
| Mean absolute value ($10^{-3}$) | 129 ± 122 | 129 ± 122 | 141 ± 131 | 143 ± 133 |
| MSE ($10^{-3}$) | 7.16 ± 11.3 | 6.96 ± 11.3 | 8.72 ± 17.7 | 9.04 ± 17.3 |
| Ratio (%) | 4.46 ± 4.67 | 4.34 ± 4.43 | 4.78 ± 5.86 | 4.84 ± 5.69 |

The linear correlations between the original and reconstructed signals were calculated for the test dataset for both the EEG and fMEG signals. The cross-correlation (Figure 4) showed a highly significant correlation between the original and reconstructed EEG and MEG signals, 0.91 ± 0.05 (p < 0.001) and 0.89 ± 0.08 (p < 0.001), respectively, at 0 lag.

**Figure 4.**
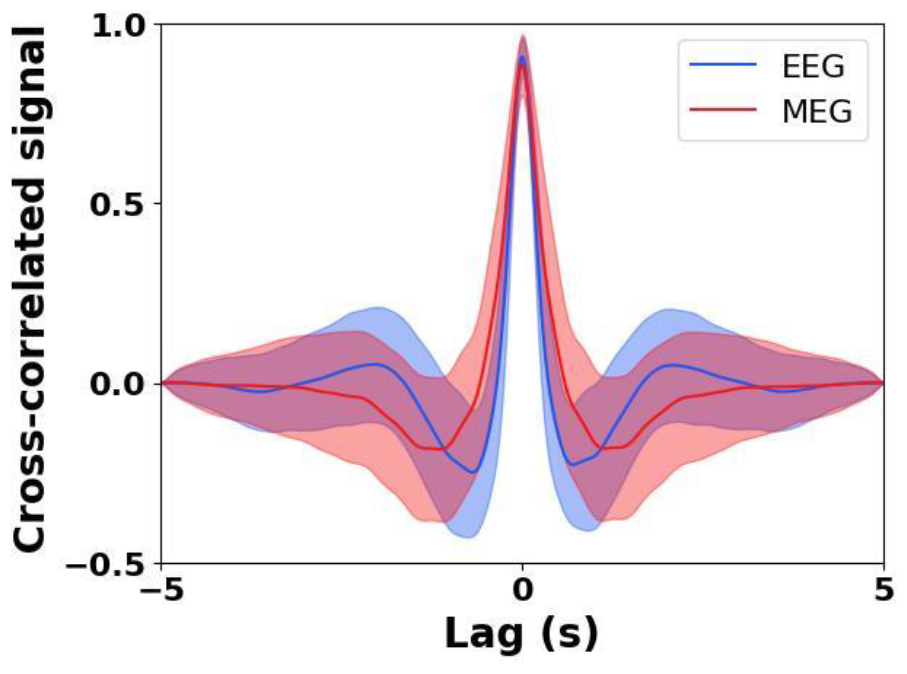
Correlation between the original and reconstructed signals. The solid line corresponds to the average and the shaded area to the standard deviation. The EEG signal is plotted in blue and the fMEG signal in red.

The frequency content of the reconstructed signals was evaluated and compared to that of the original signal for the test subset (Figure 5). CycleGAN modified the spectral content of the reconstructed signal compared to the original signal. More precisely, for the EEG signals, the spectral power of the reconstructed signal was significantly lower than that of the original in the theta (t = -6.11, p < 0.001) and alpha (t = -4.35, p < 0.001) bands (Figure 5A and 5C). However, the spectral power of the reconstructed EEG signal was not significantly different in the delta (t = 0.21, p = 0.84) or low beta (t = 1.2, p = 0.095) bands. For the fMEG signals, there were no significant differences in the spectral power (Figure 5B and Figure 5.D) between the original and reconstructed signals for any of the frequency bands: delta (t = -0.25, p = 0.80), theta (t = 0.10, p = 0.92), alpha (t = 1.23, p = 0.22), and low beta (t = 1.19, p = 0.24).

**Figure 5.**
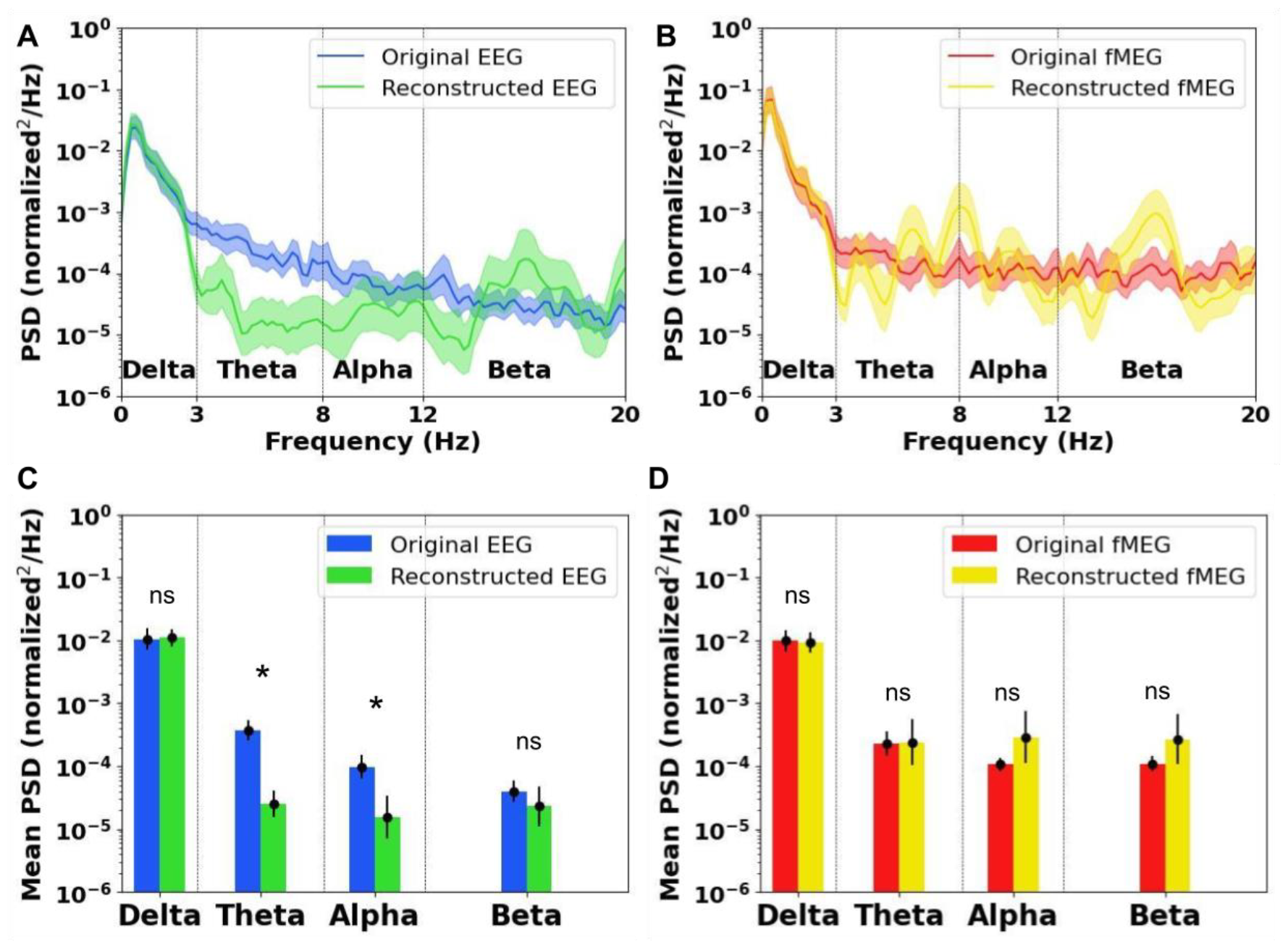
Spectral power of EEG and fMEG signals (original EEG in blue, reconstructed EEG in green, original fMEG in red, reconstructed fMEG in yellow). ★ indicates a significant difference and ‘ns’ indicates no significant difference. A) Spectral power of EEG signals, and B) Spectral power of fMEG signals. The solid lines correspond to the average and the shaded areas to the standard deviation. C) Mean spectral power over the frequency bands for EEG, and D) fMEG.

### Evaluation of fMEG signals generated from EEG

As the brain activity of the newborn is also characterized by specific features such as DB and FT, it is interesting to evaluate the abilities of CycleGAN in the EEG-to-MEG transfer of these two specific types of activities. This was carried out here only through visual inspection over a small number of events. A sample event corresponding to DB and a sample event corresponding to FT are shown in Figure 6. For the DB event, the CycleGAN transfer function replicated the slow dynamics of the EEG signal in the fMEG signal, whereas the relatively faster nested oscillations were modulated in the generated fMEG signals. For the FT, the transform kept the slow oscillation intact. Visual inspection of the results suggests that for both examples, the transform added high frequency oscillations of small energy to the fMEG signal.

**Figure 6.**
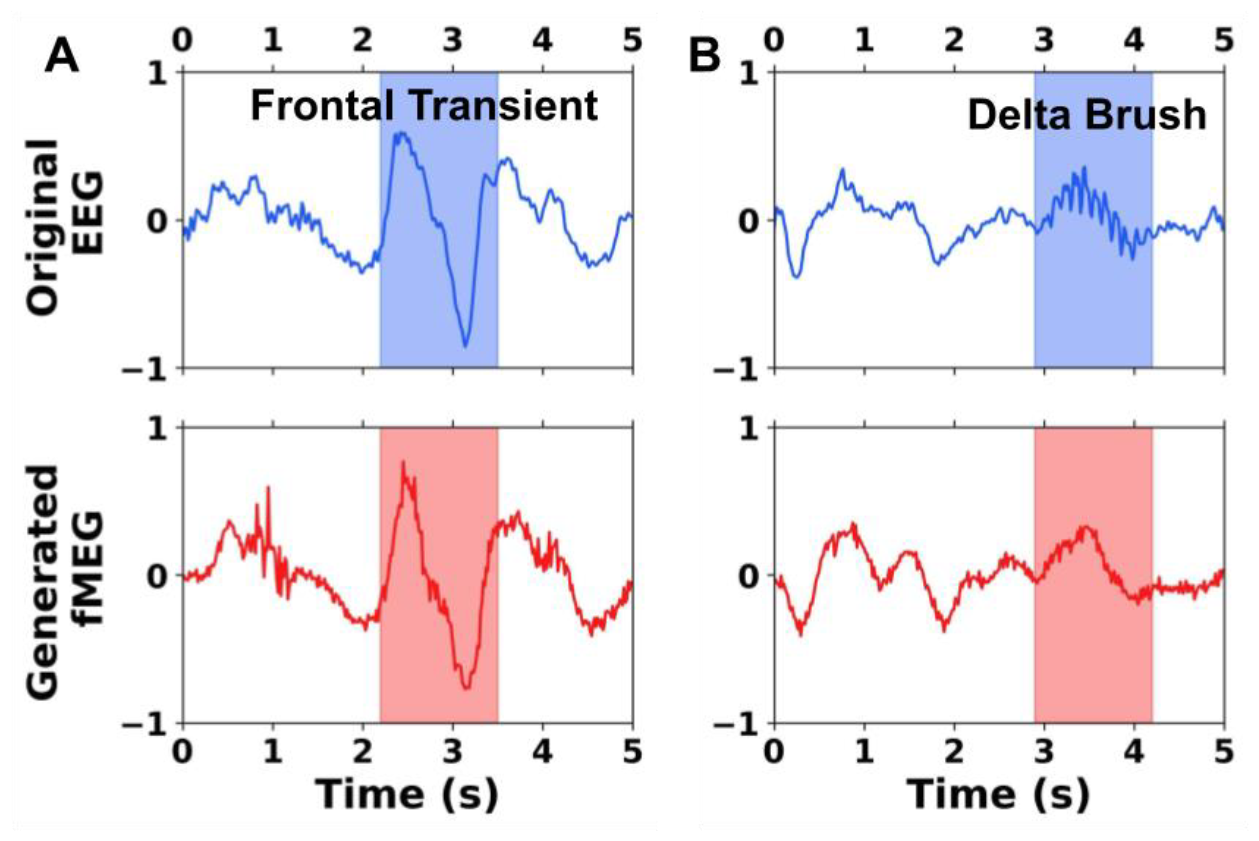
Examples of the transformation of neurobiomarkers from EEG to fMEG. A) FT transformation. B) DB transformation. The original EEG segment is shown in blue (TOP) and the generated fMEG segment is shown in red (BOTTOM). The neurobiomarkers are indicated by the shaded areas.

For further evaluation of the CycleGAN-based generation of fMEG signals, we considered the power spectra of the original and generated fMEG signals for the test data subset. Visual inspection of the spectral power (Figure 7A), as well as the mean spectral power (Figure 7B), revealed a close resemblance in the slow frequency content, corresponding to the delta band, between the original and generated fMEG signals. However, the CycleGAN model modified the higher frequency contents in theta, alpha, and beta bands. This observation fits with the induced high frequency activity observed over the generated MEG signals, as well as the modulation of the fast oscillation nested in the DB in Figure 6. Two-sample two-tailed t-tests showed that there was no significant difference between the spectral content of the original fMEG and generated fMEG signals over the delta band (t = 1.24, p = 0.22). However, the spectral content of the generated fMEG signal was significantly higher than that of the original fMEG signal in the theta (t = 2.58, p = 0.011), alpha (t = 3.33, p = 0.0012), and low beta (t = 3.66, p < 0.001) bands.

**Figure 7.**
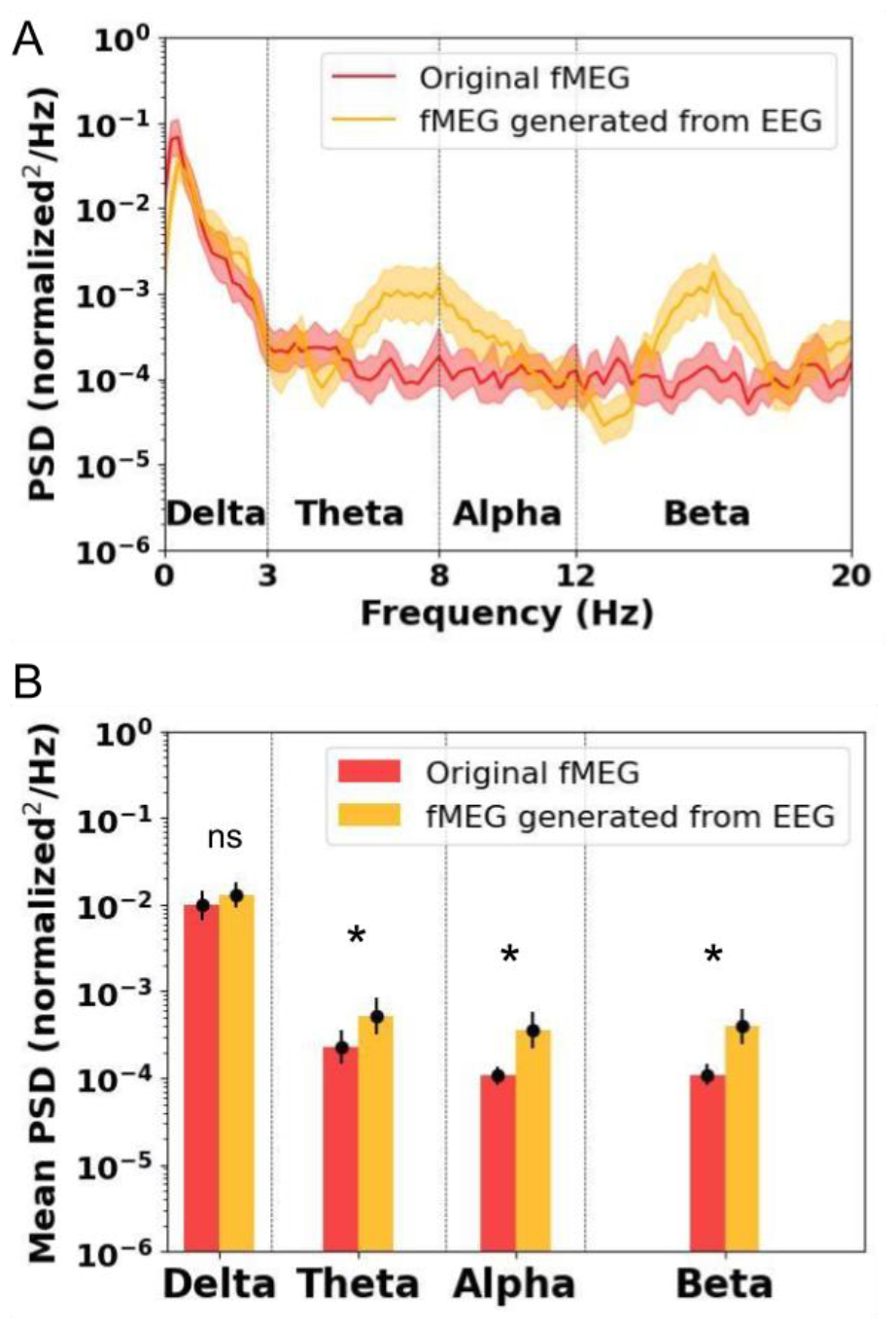
A) Spectra of fMEG signals (original in red, CycleGAN based generated in orange). The solid line corresponds to the average and the shaded area to the standard deviation. B) Mean spectral power over different frequency bands. ★ indicates a significant difference and ‘ns’ indicates no significant difference.

## Discussion

In this study, we developed a CycleGAN-based model to transform premature EEG to fMEG signals. We generated prototypical fMEG manifestations of spontaneous neural activity by creating a priori representations derived from premature EEG recordings. In general, the performance of CycleGAN was better in transforming the low-frequency content of the EEG signals into fMEG signals, suggesting more reliable evaluation of the low-frequency content in the generated fMEG signals.

The fMEG is the only tool that allows investigation of the function and dysfunction of immature neuronal networks of the fetus with high temporal resolution. This is crucial as it addresses public health concerns and has the potential to help in the identification and management of high-risk pregnancies. Such risks include prematurity, in utero asphyxia, preeclampsia, fetal growth retardation, exposure to neurodevelopmental toxins, fetal cerebral suffering, and other pathologies/causes likely to impact the dynamics of fetal cerebral functioning [47]. The prenatal assessment of neural activity necessitates the identification of fetal neurobiomarkers of neurodevelopment, similar to what is carried out using EEG in premature newborns as part of clinical routine, aimed at evaluating the normality of brain function at different periods of development and predicting neurodevelopmental outcomes [1, 48]. Our proposed approach represents the first steps toward this ambitious goal by transferring the clinical knowledge acquired using EEG in premature neonates to fMEG, before identifying the neurobiomarkers of brain development in the fetus by MEG.

### CycleGAN: an approach for unpaired signal transfer

CycleGAN is a novel generative neural network method mainly developed for image transformation, such as horse to zebra or photo to painting [30, 46]. However, it has also been applied to signals, for instance in voice conversion [49] or in the medical field for fetal ECG extraction [33]. A general challenge with generative techniques is the evaluation of the quality of the generated data [50]. Our evaluation of the transformation of EEG signals to fMEG signals and vice versa using CycleGAN showed good performance in the temporal domain, with similar MSEs and high correlations between the original and reconstructed signals for both EEG and fMEG. However, in the frequency domain, CycleGAN showed better performance in the reconstruction of low-frequency content. Indeed, the high frequencies, which lower energy in the neural signals, visible also in the power spectra, were modified in the reconstructed signals. We have two hypotheses about the origin of this modulation. One reason behind this modulation might be related to the characteristics of the CycleGAN based models. In line with this hypothesis, evidence suggests that CycleGAN does not completely preserve the color composition in image reconstruction, as it favors the dominant colors [30]. In our case, the spectral content of the neural data is determined by the temporal fractal and nested oscillations that manifest in the 1/f power law function form of the spectra [51]. This suggests that the energy of the spectral content decreases with an increase in frequency. The observed results suggest that CycleGAN gave more weight to the low spectral content of the signal in the cost functions to reduce the prediction error, as shown by the MSE. This is also visible in the high correlation values between the original and predicted signals, which again is driven by the high amplitude, low spectral content of the signals. This probably indicates an instance of mode collapse, a well-known drawback of GANs [52]. Briefly, this means that due to the adversarial nature of GAN training, the generators may specialize themself on reproducing certain aspects, named “modes” (here, the low frequency content) of the training data and not others “mode collapse” (here, the high frequency content) [53, 54]. In CycleGAN, we employed a linear combination of the losses with the same weight. However, each part can have a different impact and it can be necessary to define different weights manually or automatically [55]. In addition, to overcome this issue in future studies and with the aim to create better predictions of high spectral content, it might be possible to develop parallel models to predict the low and high spectral content separately with a mixing unit for weighted superposition of the outputs following a power law function. Another reason behind the observed modulation of the high frequency content might be related to the nature of the fMEG signals. As depicted in Figure 5, the fMEG signals show less conformity to the 1/f power law form observed in the spectrum of EEG signals, and instead exhibit a relative increase in the high frequency power compared to EEG signals. This might lead to the observed modulations in the results; precisely, that the EEG to fMEG transformation adds to the high frequency content of the signal, and that the fMEG to EEG transformation reduces the high frequency content.

### Transfer of neurobiomarkers from EEG to fMEG

The principal interest of EEG/fMEG transfer is the visualization of fMEG signals, to provide insights of what the early aspects of fetal spontaneous activity recorded by MEG would look like. In this study, we focused on two particular endogenous neurobiomarkers that are present at this gestational age: DB and FT [1]. The slow waves of the FT and DB had similar forms in the original EEG and generated fMEG. However, the DB lost their fast nested activity in the transform. This suggests that without knowing the transformation of fast activities, it may be preferable to focus on low-frequency content (the slow waves) in future automatic algorithms for fMEG detection of these neurobiomarkers.

## Conclusion

Our CycleGAN-based model offers the opportunity to transform premature EEG into fMEG signals and allows visualization of fMEG representations of various EEG templates and types of activity, including those used in the clinical applications for the evaluation of neurodevelopment. It provided a good representation of the low-frequency content of the signals but modified the high-frequency content. This aspect is important for the study of neurobiomarkers, especially DB. Moreover, by extending this study to younger premature infants and fetuses, the same issue is likely to occur for theta temporal slow-wave activity, in which fast nested activities are locked to the trough of the delta wave [21]. This modification of high-frequency signals is related to the tendency of GANs toward mode collapse. Recent approaches, such as diffusion [56], are known to mitigate mode collapse. Therefore, a promising research direction would be to extend our model with the application of diffusion models instead of GANs.

## Data Availability

All data produced in the present study are available upon reasonable request to the authors

## Acknowledgments

The authors are grateful to the Amiens University Hospital EEG technicians and Tübingen University Hospital MEG technicians for data acquisition. We thank the parents and families who consented to take part in the study.

